# Feasibility of Electroencephalography-Based Detection of Single-Flash Microperimetry Stimuli: A Proof-of-Concept Study

**DOI:** 10.1101/2025.09.12.25335536

**Authors:** Muhammad Najam Dar, Antonio Neto Severo de Castro, Zoha Zahid Fazal, Kholood Janjua, Mohammed Abdul Saqhlain Shaik, Tehmina Sheharyar, Mohamed Ibrahim Ahmed, Yasir Jamal Sepah

## Abstract

**Purpose:** To evaluate the feasibility of detecting single-trial cortical responses to individual microperimetry (MP) stimuli using electroencephalography (EEG) under non-ideal synchronization conditions, and to explore EEG-based stimulus registration independent of patient responses.

**Methods:** This proof-of-concept study acquired EEG data from two healthy participants (12 trials) during MP testing using low- and high-intensity single-flash stimuli. Occipital EEG signals were recorded with an 8-channel portable system, band-pass filtered (4–49 Hz), normalized, and segmented into 600-ms epochs time-locked to MP stimuli using offline synchronization, resulting in an estimated temporal uncertainty of ∼250 ms. A bidirectional long short-term memory (BiLSTM) deep learning model classified stimulus-present versus stimulus-absent EEG segments. Performance was assessed using accuracy, sensitivity, specificity, and F1-score, with emphasis on feasibility rather than generalizability.

**Results:** Across trials, EEG-based classification performance exceeded chance levels. For high-intensity stimuli, detection accuracy approached 80%, while low-intensity stimuli demonstrated greater inter-trial variability. Occipital electrode configurations consistently outperformed parietal or combined montages, consistent with visual cortex neuroanatomy. Despite the absence of hardware-level synchronization and single-trial analysis, detectable neural signatures of isolated MP flashes were observed.

**Conclusion:** These findings demonstrate the feasibility of detecting single-flash MP stimuli from occipital EEG using deep learning, even under constrained acquisition conditions. While not intended for immediate clinical deployment, this work motivates future studies incorporating precise synchronization, optimized stimulus designs, and larger cohorts to evaluate EEG-augmented microperimetry as a potential objective adjunct to subjective functional testing.

**Highlights:**

- EEG detects single-flash microperimetry stimuli without hardware-level synchronization
- Occipital EEG channels enable stimulus detection independent of patient responses
- BiLSTM deep learning decodes non-repetitive, long-duration microperimetry flashes
- Detection accuracy approaches 80% for high-intensity microperimetry stimuli

## Introduction

Microperimetry (MP) enables localized assessment of macular function by mapping retinal sensitivity while accounting for fixation stability. It is widely used in clinical trials and longitudinal monitoring of maculopathies such as age-related macular degeneration (AMD) and diabetic macular edema (DME), where it can reveal subtle functional impairments not captured by conventional acuity testing. MP can also detect early functional loss in para-atrophic regions of geographic atrophy (GA) and Stargardt disease (SD) before central vision loss occurs. By bridging the gap between structural and functional assessments, MP facilitates earlier detection of macular dysfunction and supports timely therapeutic decision-making for the more than 200 million individuals affected by retinal degenerative diseases worldwide^1,2^.

Despite its clinical relevance, MP remains highly dependent on patient attention, fatigue tolerance, and subjective stimulus reporting, which can compromise test reliability—particularly in elderly, pediatric, or cognitively impaired populations^3^—thereby limiting its utility in clinical trial settings. Cognitive decline, testing fatigue, inattentiveness or non-cooperation, infrequent physiological scotoma assessment, and response variability are well-recognized challenges in geriatric MP testing^4–6^. Although MP has shown promise in pediatric ophthalmology for monitoring hereditary retinal diseases^7^, children similarly demonstrate poorer test–retest reliability, lower mean sensitivity, and reduced fixation stability compared with adults^8^. Collectively, these limitations underscore the need for more objective approaches to stimulus registration in MP.

Several objective perimetry methods have been developed, each with distinct advantages and limitations. Multifocal pupillographic objective perimetry (mfPOP) enables rapid, binocular testing with demonstrated clinical utility in macular and optic nerve disease^9,10^. Multifocal visual evoked potential (mfVEP) models further provide objective cortical readouts using scaled stimuli^11–13^. However, these approaches typically require specialized hardware, proprietary stimulus designs, and optimized acquisition environments, which may limit their integration into existing clinical MP workflows. Similarly, focal electroretinography (FERG) has demonstrated improved sensitivity for detecting macular dysfunction in intermediate AMD compared with MP, although differences in patient demographics and limited sample size of the study may confound interpretation^14^.

The present proof-of-concept introduces a paradigm that leverages patient-independent, long-duration MP stimuli to evoke electroencephalography (EEG) responses, in contrast to traditional VEP paradigms that rely on flickering or patterned stimuli. While VEPs recorded using head-mounted EEG devices have outperformed standard automated perimetry in glaucoma detection¹C, and abnormal VEP latencies or amplitudes can reveal visual pathway disorders such as in multiple sclerosis^13^, the isolated and prolonged nature of MP stimuli renders classic time-locked VEP waveforms difficult to detect in single trials. Accordingly, we adopted a bidirectional long short-term memory (BiLSTM)–based model to decode non-repetitive, variable-intensity MP stimuli and to assess stimulus detectability of different EEG electrode configurations without relying on waveform averaging or explicit peak identification^11,15^.

Importantly, the goal of this study was not to propose MP stimuli as optimal for EEG-based perimetry, nor to compete with established mfVEP or mfPOP methodologies. Rather, we sought to evaluate whether meaningful neural information can be extracted from the constrained stimulus design and timing limitations imposed by currently available clinical MP devices. Given the widespread adoption of MP as a functional endpoint in retinal clinical trials, exploring objective augmentation within this constrained framework represents a relevant translational question and forms the basis of this feasibility investigation.

## Methods

### Data Acquisition

We recorded EEG signals with timestamps corresponding to the presented MP stimuli with a 250 Hz sampling frequency. We used the Nidek-MP1 device^16^ for its custom settings and enabled fixation monitoring although Goldmann IV stimuli in Nidek-MP1 are suprathreshold relative to Goldmann III norms. The low-intensity condition should hence be interpreted as relatively dim rather than near-threshold, and comparisons to standard perimetric thresholds should be made cautiously. We used an OpenBCI complete ultracortex 8-channel EEG headset^17^ with a cyton bio-amplifier board for signal acquisition. We adjusted EEG electrode locations to capture the response primarily from the visual cortex with parietal (P3, P4), parieto-occipital (PO3, PO4, POz), and occipital (O1, O2, Oz) channels^18^ as shown in **Figure 1**. OpenBCI graphical user interface was used for EEG signal recordings in a portable document format (PDF) for offline processing.

**Figure 1:**
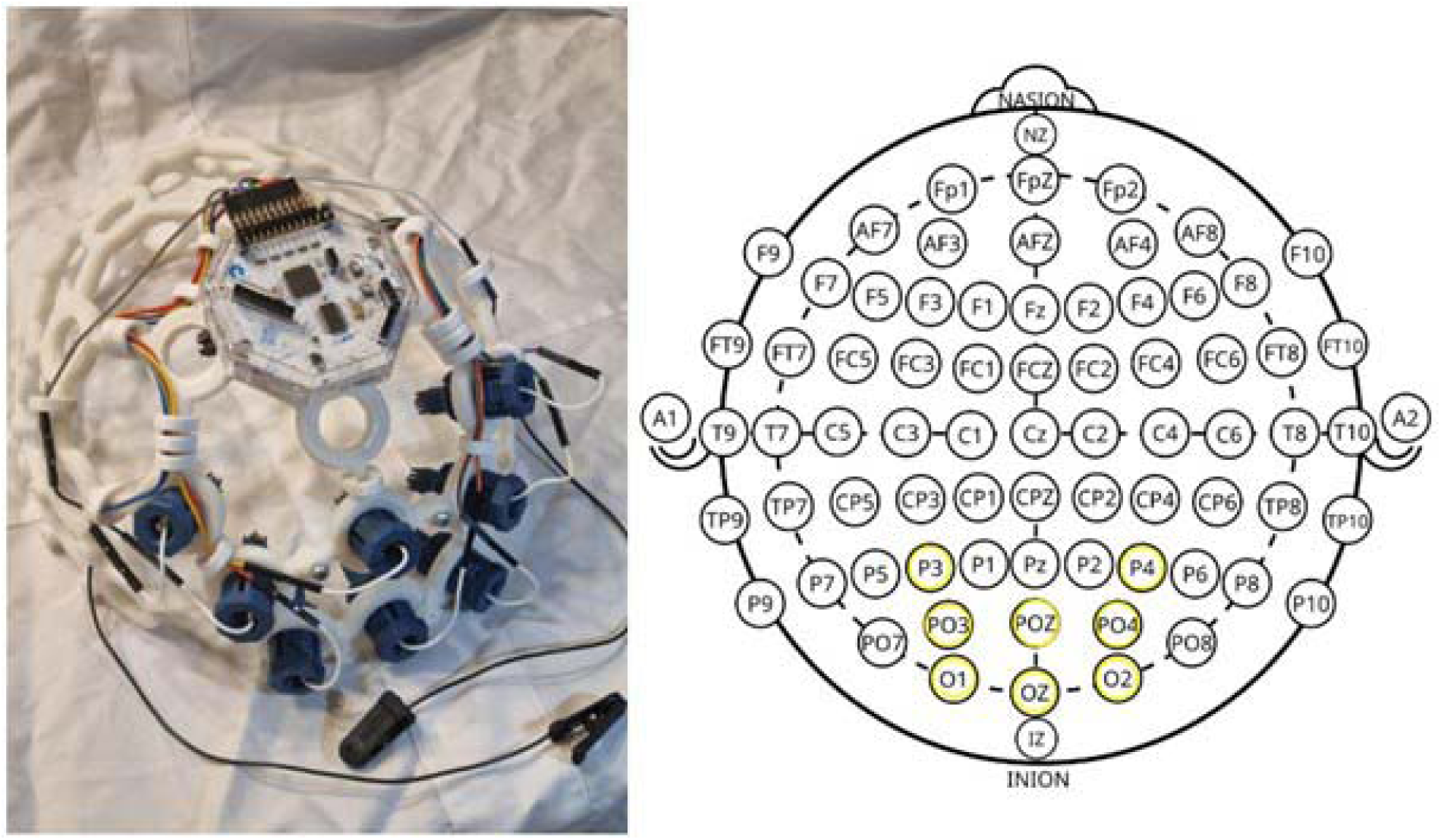
Customized channel configuration to capture the visual cortex response to flash stimulus of microperimetry.

Nidek-MP1 luminance ranges from is 0 dB (highest) to 20 dB (1.27 cd/m2), which is the lowest and equals background luminance. The trials were divided into two settings featuring high- and low-intensity stimuli. During high-intensity settings, we used either 0 dB (127 cd/m2) or 4 dB (50.6 cd/m2) stimulus luminance while 16 dB (3.2 cd/m2) was used for low-intensity stimulus settings. This was because 16 dB is slightly above the healthy macular sensitivity of 18.33 ± 2 dB for Nidek-MP1^16^, and historic MP protocols show that points missing 16 dB stimulus threshold reliably indicate emerging scotomas before visual acuity loss^19^. Moreover, test–retest data in the GA cohort show that a change of ± 4 dB in point-wise sensitivity reliably exceeds measurement variability, making 4 dB a meaningful near-floor benchmark for true progression versus noise^20^. All trials were performed using the right eye, while the left eye was covered during the exam. For synchronization, MP exams and EEG recording using the OpenBCI GUI were started at the same time. For stimulus timestamp synchronization, MP screen videos were recorded, showing the stimulus onset, stimulus offset, and patient response timestamp of pressing the trigger for use as ground truths for event-related potentials, as shown in **Figure 2**.

**Figure 2:**
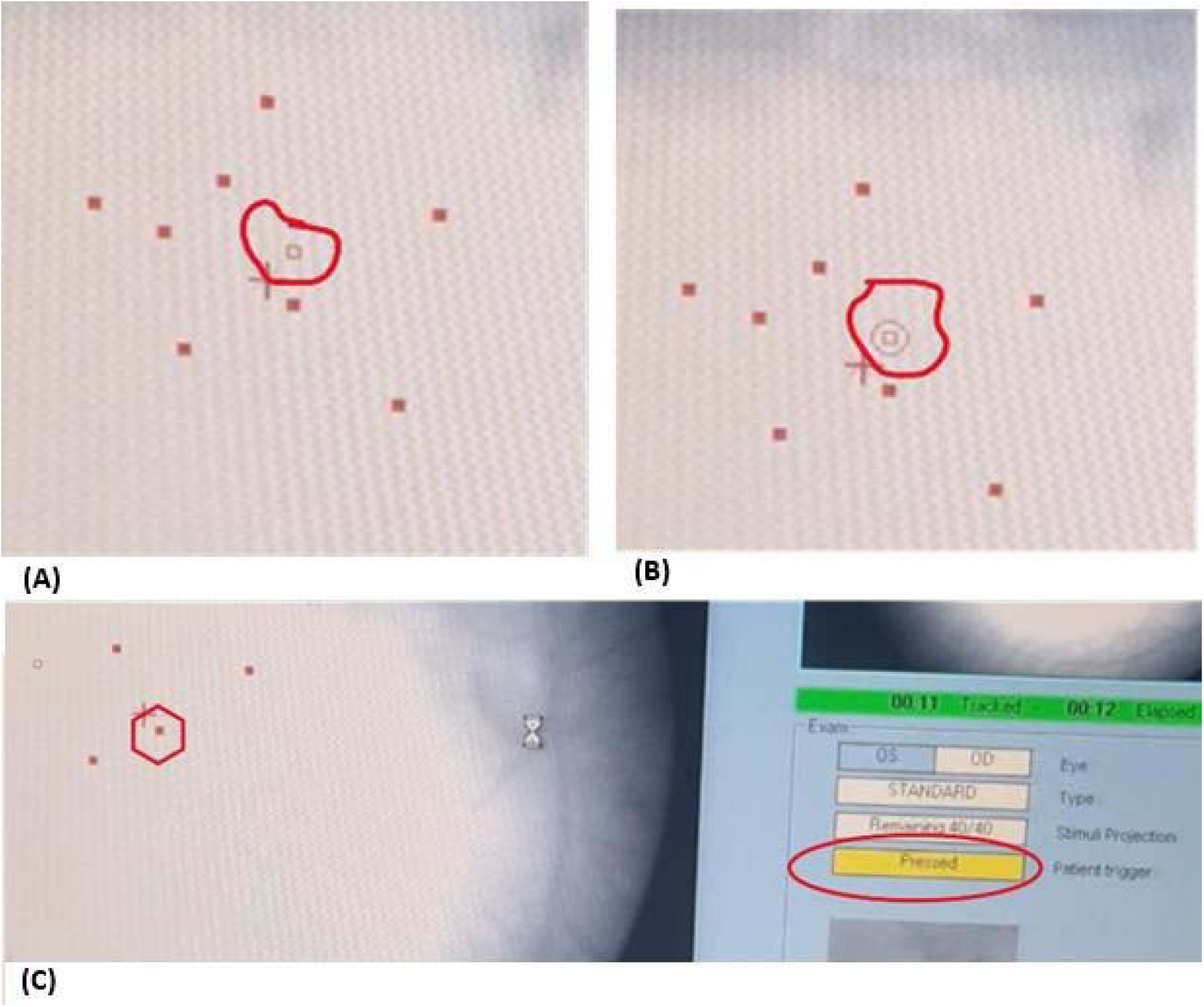
MP recorded video screenshots depicting timestamp extraction process using DataVyu software. **(A)** The hollow square shows target next location of stimulus to be presented to patient’s retina **(B)** Stimulus onset timestamp when a circle appears enclosing the hollow square **(C)** Stimulus offset timestamp when outer circle disappears and square filled with red color, patient trigger will be shown pressed and separately logged as patient response timestamp.

Standard MP devices lack the ability to precisely time-lock MP stimulus with data streaming of external devices using hardware-based synchronization. Future implementations will incorporate hardware-based triggering (e.g., photodiode-based onset detection or Open Perimetry Interface-compatible systems) to reduce timing uncertainty to the millisecond range. For this pilot study, as illustrated in **Figure 3**, we performed offline synchronization of the MP stimuli with EEG signals by reviewing the MP screen video recording in slow motion (¼ of the original speed) using the DataVyu software^21^ and exporting the timestamps in Microsoft Excel format for each trial. The approximate timing error introduced by this manual approach of synchronization is 250 ms.

**Figure 3:**
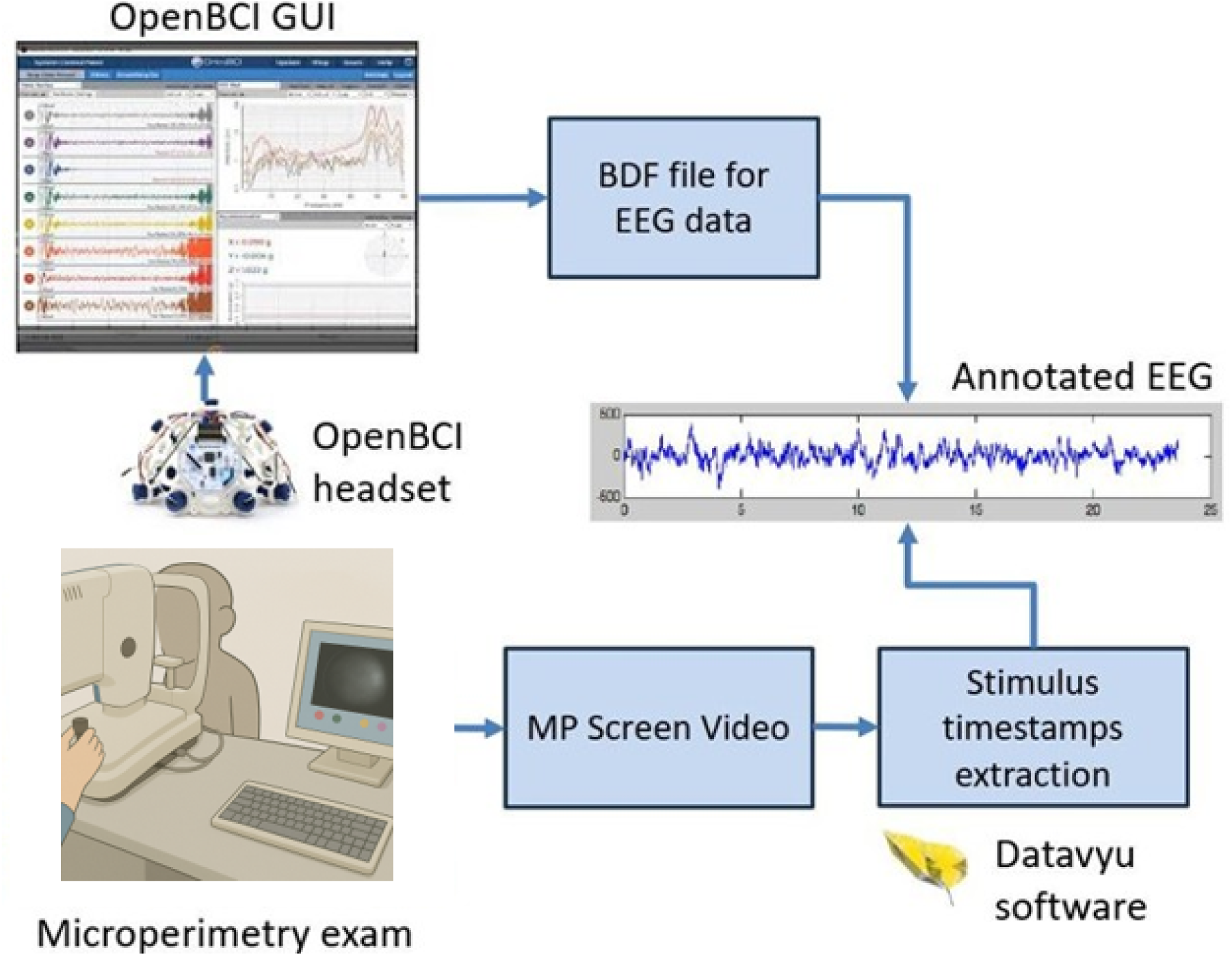
Flow chart for EEG data acquisition during microperimetry examination, and EEG data annotation for stimulus timestamps.

### Study Participants

This pilot study incorporated data from 12 trials conducted on two subjects with normal visual functions to validate the hypothesis that advanced machine learning techniques can elicit neural responses of non-flickering and long-duration MP stimulus. The small sample size was intentionally chosen for feasibility testing and is not intended to support claims of inter-subject generalizability. Informed consent was obtained from both participants who were guided about the use and expected outcomes of this study. The first subject (ID-1) is a male in his early 30s with best-corrected visual acuity (BCVA) of 20/20, while the second subject (ID-2) is a female in her early 20s with BCVA of 20/15. Both study participants were tested without any corrective lenses or medications used during trials. Five trials were conducted with high-intensity and seven trials were conducted with low-intensity stimulus settings. The eye laterality, length of each trial, fixation stability rate, and number of stimuli are presented in **Table 1**.

**Table 1:**
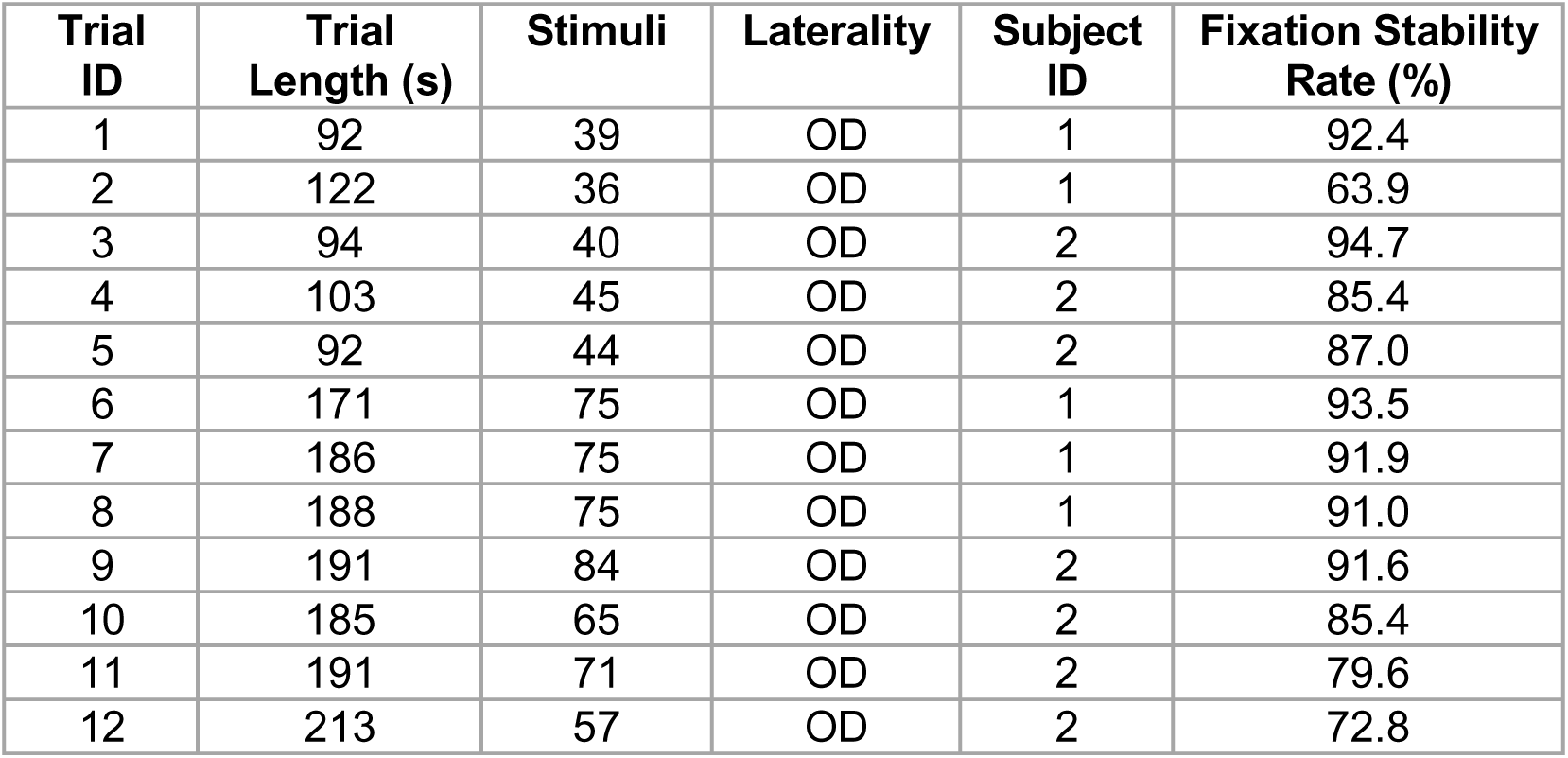
Details of data acquired including laterality, length of trial, fixation stability rate, and number of stimuli presented for each trial.

| Trial ID | Trial Length (s) | Stimuli | Laterality | Subject ID | Fixation Stability Rate (%) |
| --- | --- | --- | --- | --- | --- |
| 1 | 92 | 39 | OD | 1 | 92.4 |
| 2 | 122 | 36 | OD | 1 | 63.9 |
| 3 | 94 | 40 | OD | 2 | 94.7 |
| 4 | 103 | 45 | OD | 2 | 85.4 |
| 5 | 92 | 44 | OD | 2 | 87.0 |
| 6 | 171 | 75 | OD | 1 | 93.5 |
| 7 | 186 | 75 | OD | 1 | 91.9 |
| 8 | 188 | 75 | OD | 1 | 91.0 |
| 9 | 191 | 84 | OD | 2 | 91.6 |
| 10 | 185 | 65 | OD | 2 | 85.4 |
| 11 | 191 | 71 | OD | 2 | 79.6 |
| 12 | 213 | 57 | OD | 2 | 72.8 |

### EEG Signal Processing

The EEG pre-processing was used to prepare the signal for a deep-learning model to classify stimulus and non-stimulus segments. A limited eight-channel montage was selected to evaluate minimal EEG configurations compatible with low-cost and portable systems, rather than to replicate dense mfVEP electrode arrays. This design choice prioritizes feasibility and accessibility over spatial resolution and is not intended to replace established mfVEP methodologies. The detailed framework is presented in **Figure 4**. EEG data were re-referenced using the common median reference (CMR) as a pre-processing step to improve signal quality. The CMR re-referencing technique is less sensitive to outliers, highlights local neural activity by removing global fluctuations, environmental and muscle noise, and works well even when some channels are corrupted. The median of all eight channels was computed and subtracted from each channel. Suppose Si(t) is the EEG signal from channel i at time t, and M(t) represents the median across all eight channels, the CMR is represented by **equation 1**.

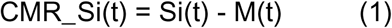

**Figure 4:**
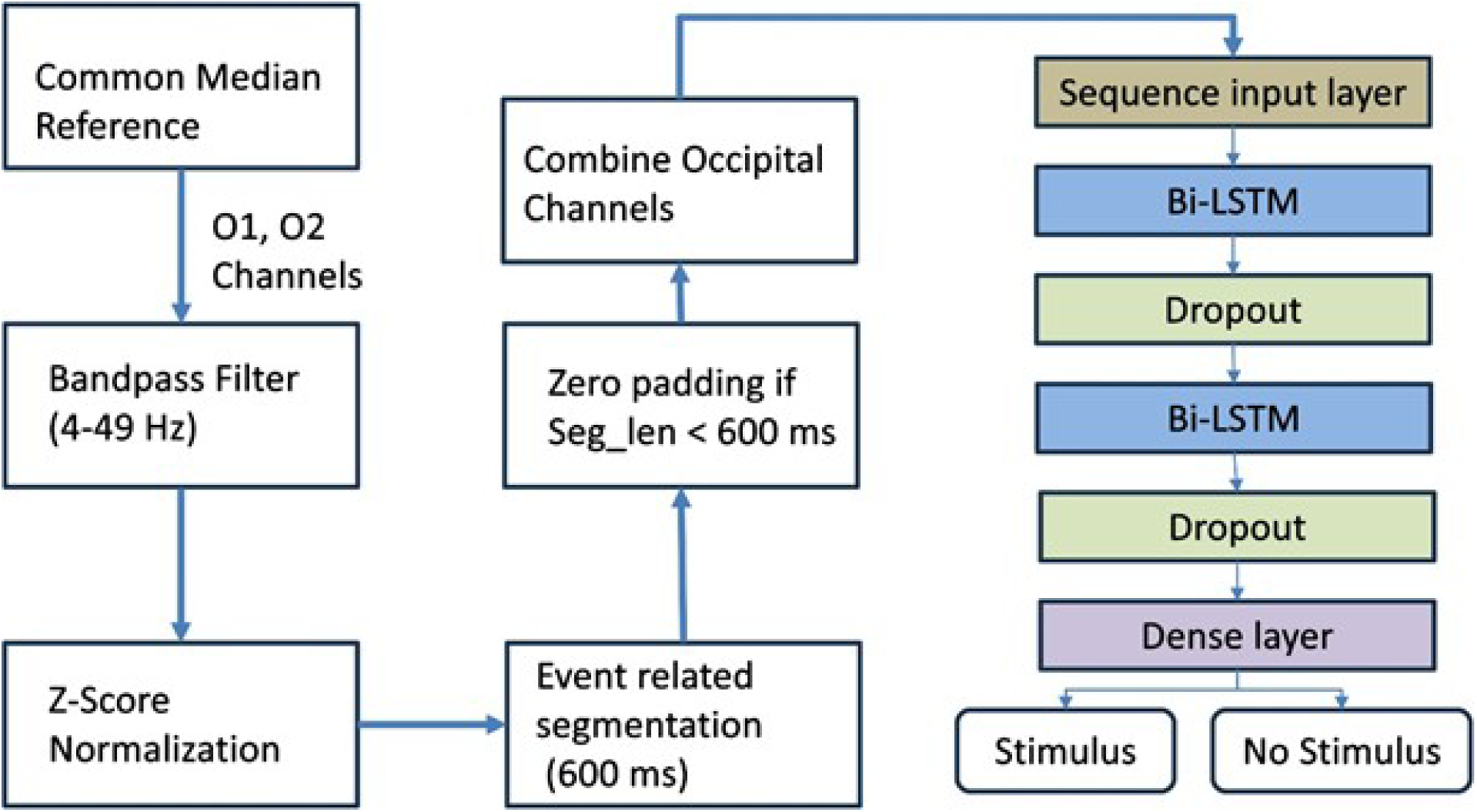
Flow chart for detection of stimulus and non-stimulus EEG segments using O1 and O2 channels.

The artifact removal and standardization of the EEG signal is significant for uniform scaling of features to test our hypothesis of exploring deep learning architecture for subtle response elicitation of non-standard visual stimuli. To achieve this, we performed further artifact removal, and z-score based data normalization. After CMR, the slow drifts in EEG signals obscuring P1 and N1 peaks were eliminated by using the low cutoff frequency of 4 Hz. Similarly, the muscle and electrical interference noise was removed by using the high cutoff frequency of 49 Hz. To achieve this, a bandpass Butterworth filter of 4-49 Hz is designed with filter order 4 to remove both low and high-frequency noises. This bandpass filtering helps optimize the EEG signal for flash signal analysis on neuronal visual processing for parieto-occipital electrodes. After bandpass filtering, all channels were normalized using z-score normalization. For that purpose, the average of the signal is subtracted and then divided by the standard deviation to extract uniform scale features, required as input for a deep-learning architecture. In the absence of built-in hardware synchronization capabilities within the MP device, we adopted a software-based preprocessing approach to approximate temporal correspondence between visual stimuli and neural responses. To achieve this, we performed event-related segmentation on the normalized EEG data based on stimulus timestamps, as shown in **Figure 5**.

**Figure 5:**
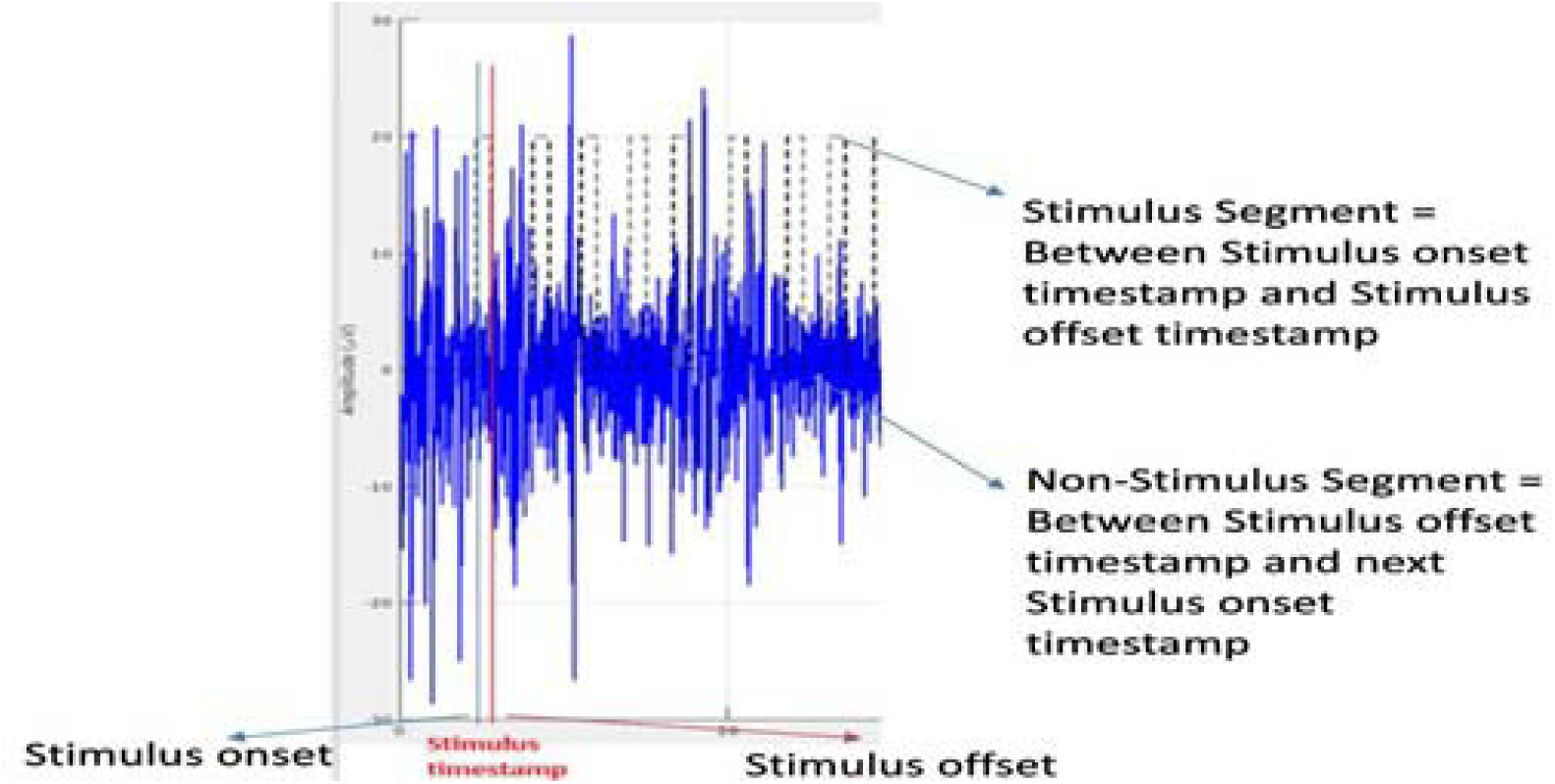
The O1 channel of EEG signal depicting stimulus timestamp, and segmentation of 600 ms chunk before the stimulus timestamp as stimulus segment and everywhere else as non-stimulus segment.

The segmentation window of 600 ms before each stimulus offset timestamp is considered a stimulus segment. The rationale for choosing 600 ms pre-offset window is to consistently capture both the full stimulus duration, preceding early onset neural activity significant for assessing baseline dynamics, while avoiding inclusion of post-perceptual or motor-related processes. Secondly, the offset timestamp is captured using DataVyu software displaying recorded video of the MP screen; therefore, there is a time buffer between the actual stimulus offset, the patient triggering the push button, and the results displayed on the recorded video of the MP screen. The VEP response to a flash stimulus is normally 300 ms, but to cater to the potential lag, a 600 ms window is used in our methods. The other part of the signal outside the stimulus segment is considered a non-stimulus part, which is further divided into 600-ms segments to make all the segments equal in length. Zero padding was performed if any segment lengths were less than 600 ms. This preprocessing of windowing larger lengths was necessitated to ensure meaningful temporal analysis in the absence of hardware-level time-locking.

### Deep Learning Model

The context of isolated single-flash, long-duration MP stimuli poses unique challenges for binary classification between the stimulus and the non-stimulus EEG segments, which necessitates the use of a more temporally expressive architecture. The single-flash and relatively long-duration of the MP device stimulus results in inherently low signal-to-noise ratio (SNR) due to the absence of repetitive averaging and temporal dispersion of neural responses. The temporal dispersion is further complicated with the lack of native support of MP devices for hardware-level synchronization, which requires segmentation windowing of a length (600 ms) larger than the normal VEP pattern. This increased window size introduces the risk of overlapping noise and unrelated brain dynamics, making conventional feedforward or shallow recurrent models insufficient for discriminating subtle, temporally distributed signal patterns.

To address these MP-based methodological constraints, we employ a stacked BiLSTM architecture. The model is configured with two sequential BiLSTM layers, each followed by a dropout layer. The dual BiLSTM layers capture temporal dependencies and contextual features from past and future states by processing input sequences in both forward and backward directions. The abstract-level temporal features were extracted by the first BiLSTM layer, while deeper-level patterns were extracted in the second BiLSTM layer. The dropout layer helps mitigate overfitting by randomly deactivating neurons during model training. The proposed architecture offers the ability to model long-range temporal dependencies within an extended EEG window length and context-aware sequence learning because of the bidirectional structure of LSTM. The stacking of multiple BiLSTM layers allows the network to extract higher-level spatiotemporal features sensitive to subtle stimulus-induced neural responses while being robust to background noise and timing variability. The mini-batch size of 8 was used with an adaptive moment estimation (ADAM) optimizer during the training of this architecture.

## Results

We first report overall model performance per trial for the two stimulus intensity conditions, as shown in **Table 2**, followed by a comparison of electrode montages, as shown in **Table 3**. We then describe how stimulus intensity and electrode selection affected classification accuracy and error patterns. The experimentation protocol was defined to test the hypothesis of the ability of deep-learning architecture to elicit subtle neural responses of visual stimulus in the context of the MP exam. For each of the 12 trials, the data was split into training, validation, and testing based on several stimuli presented during the trial. Validation and test sets were 15% and the train set was 70% for each trial.

**Table 2:** Performance metrics in percentage for stimulus and non-stimulus EEG segment classification for all low intensity and high intensity stimuli trials.

| Trial ID | Intensity | Accuracy | Precision | Sensitivity (TPR) | F1-Score | Specificity (TNR) | Cohen's Kappa |
| --- | --- | --- | --- | --- | --- | --- | --- |
| 1 | High (0/4 dB) | 77.42 | 50.00 | 14.29 | 22.22 | 95.83 | 13.55 |
| 2 |  | 80.00 | 45.45 | 71.43 | 55.56 | 81.82 | 43.46 |
| 3 |  | 70.97 | 33.33 | 12.50 | 18.18 | 91.30 | 4.78 |
| 4 |  | 74.29 | 50.00 | 22.22 | 30.77 | 92.31 | 17.75 |
| 5 |  | 70.97 | 25.00 | 14.29 | 18.18 | 87.50 | 2.11 |
| 6 | Low (16 dB) | 75.44 | 50.00 | 28.57 | 36.36 | 90.70 | 22.52 |
| 7 |  | 78.69 | 62.50 | 33.33 | 43.48 | 93.48 | 31.81 |
| 8 |  | 58.06 | 31.03 | 60.00 | 40.91 | 57.45 | 13.24 |
| 9 |  | 76.19 | 57.14 | 25.00 | 34.78 | 93.62 | 22.86 |
| 10 |  | 77.42 | 46.15 | 46.15 | 46.15 | 85.71 | 31.87 |
| 11 |  | 69.84 | 14.29 | 7.14 | 9.52 | 87.76 | -6.21 |
| 12 |  | 74.29 | 35.00 | 58.33 | 43.75 | 77.59 | 28.41 |

**Table 3:**
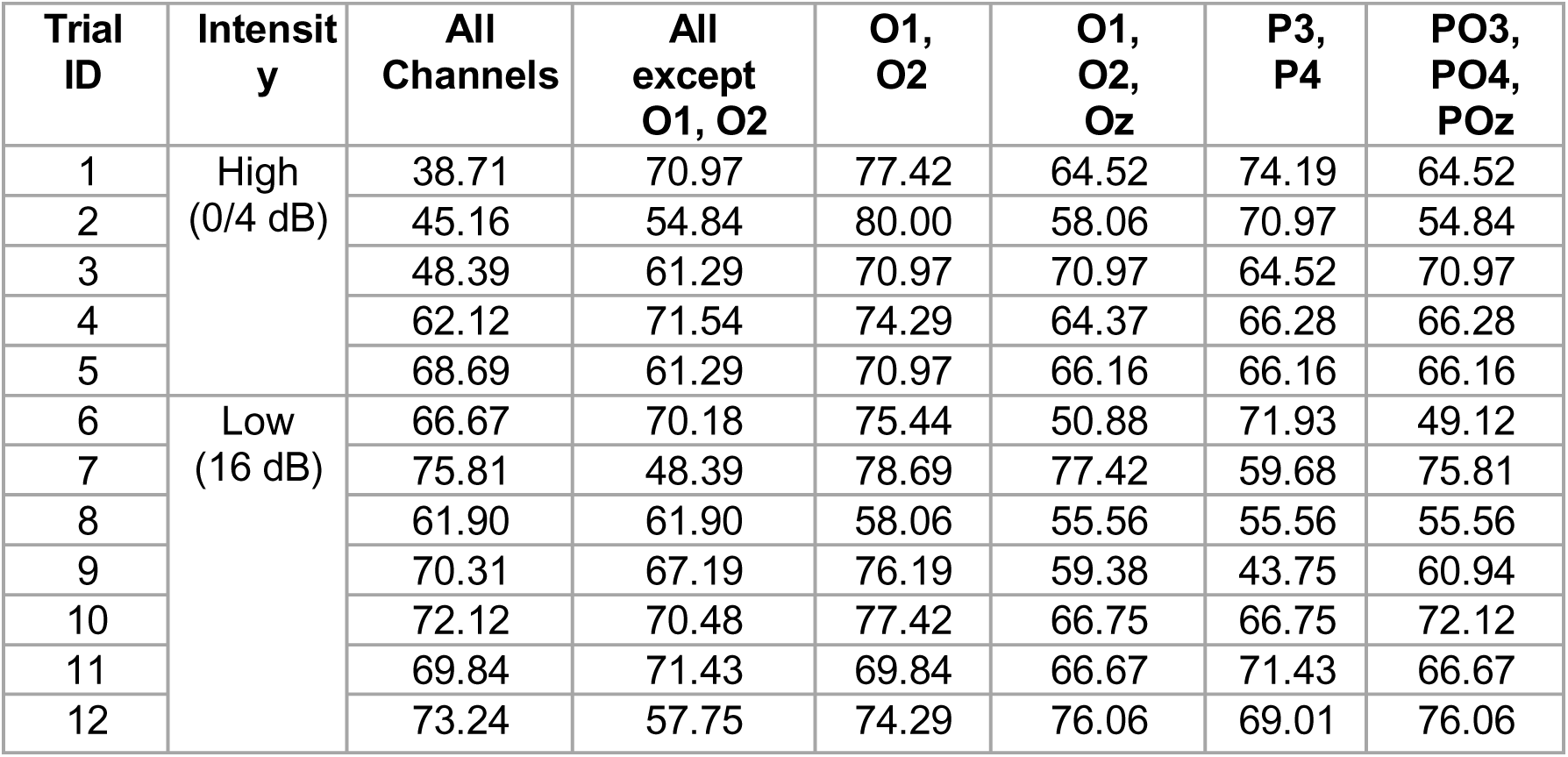
Accuracy comparison in percentage for analysis with different combinations of EEG electrodes.

Given strong class imbalance between stimulus and non-stimulus segments, accuracy alone was insufficient to characterize model performance and was therefore interpreted alongside F1-score, sensitivity, specificity, precision, and Cohen’s kappa. Accuracy measures the overall proportion of correct classification, while precision measures the proportion of stimulus predictions that are correct according to the ground truth. Specificity measures the correct classification of non-stimulus segment predictions, while the F-1 score balances precision and sensitivity. The detailed results of all the clinical trials are presented in **Table 2**. Because the sample is small, wide variability is expected, as shown in different trials for this pilot study of hypothesis testing.

### High Intensity (0/4 dB) Performance

For the high-intensity group, accuracy ranges from approximately 70.97% to 80.00%, depicting consistency in overall predictions. In one trial, a precision of 50.00% with a low sensitivity of 14.29% indicates that while half of the positive predictions are correct, many stimuli segments are misclassified. Another trial reflects a precision of 45.45% but a much higher sensitivity of 71.43%, depicting a model behaving aggressively to identify stimulus segments but at the cost of precision. Generally high (81.82% to 95.83%) values of specificity suggest that the model reliably identifies non-stimulus segments, mainly due to large proportions in the data. The large proportion of non-stimulus segments is because there is a gap of 2-3 sec between each stimulus presentation, while the stimulus only lasts for 200 ms. Cohen’s Kappa values range from about 2.11 to 43.46, indicating that despite decent accuracy, the agreement may not be robust in some scenarios. This variability in performance metrics implies that the model behavior is sensitive to class imbalances of stimulus and non-stimulus segments.

### Low Intensity (16 dB) Performance

Performance in the seven low-intensity trials was more variable (accuracy range 58% to 79%). One trial had an outlier low accuracy (58%). Precision ranged from an extremely low 14% up to 62%, and sensitivity from 7% up to 60%. This indicates some low-intensity trials were very challenging, with the model missing most stimuli (as seen in one trial with only 7% sensitivity), whereas in other trials the model achieved moderate success. Notably, even when sensitivity was higher (60%), precision tended to be moderate (31%), again reflecting the precision-sensitivity trade-off. Most Cohen’s kappa values were positive but low (several in the 20–30 range, with one trial yielding a slightly negative kappa of –6), underscoring the difficulty the model had consistently detecting faint stimuli.

In summary, the model could detect high-intensity flashes with moderate success and relatively consistent accuracy, but performance for low-intensity (dim) stimuli was more erratic and generally lower. This suggests that detecting very low luminance stimuli via EEG is feasible but prone to errors, likely due to weaker neural signals and lower signal-to-noise ratio.

### Comparison of EEG Electrode Combinations

During our experiments, we covered the parietal, parietal-occipital, and occipital regions using P3, P4, PO3, PO4, POz, O1, O2, and Oz EEG electrodes, respectively, key for visual processing. For the classification of subtle responses in EEG channels due to individual flash stimuli, we empirically tested different combinations of electrodes using the proposed methodology.

- **O1–O2 vs. Others:** In general, a combination of O1 and O2 performs better than other combinations of electrodes as described in **Table 3**. This is because the occipital electrodes are positioned right over the primary visual cortex. O1 and O2 performed better than their combination with Oz, except in one of the twelve trials. This can be due to the ability of lateral occipital electrodes to capture more focal, lateralized visual responses compared to Oz.
- **Oz and Noise:** The central location of the Oz electrode is also susceptible to more eye or muscle artifacts resulting in a low signal-to-noise ratio, reflecting a low-performance combination when added to O1 and O2.
- **Parietal channels’ Role:** These channels are primarily involved in spatial processing, attention, and integration of visual information. In some of the outliers, parietal channels performed better than occipital channels for stimulus detection. This can be due to the improvement in signal-to-noise ratio through broader coverage of visual and parietal cortices. These are occasional situations where parietal or broader parietal-occipital montages yield better flash stimulus detection than the standard occipital pair alone.
- **All channels vs. subsets:** Notably, using all 8 channels together did not produce the best results (**Table 3**), likely because the inclusion of non-visual channels added noise or irrelevant signals that confused the classifier, reinforcing the value of focusing on relevant channels.

Because only two subjects were studied, electrode performance comparisons should be interpreted as exploratory rather than definitive indicators of optimal montage selection.

## Discussion

This pilot study demonstrates that single-flash stimuli presented during a MP examination can be detected via occipital EEG using deep learning. Under optimal conditions—high-intensity stimuli and occipital electrode selection—the proposed BiLSTM model achieved classification accuracies approaching 80% for distinguishing stimulus-present from stimulus-absent EEG segments (**Table 2**). These findings support the feasibility of capturing subtle visual cortex responses to individual 200-ms flashes without trial averaging. However, detection of lower-intensity stimuli was less reliable, and overall performance varied substantially across trials, reflecting the challenging signal-to-noise conditions inherent to this experimental setup.

A key methodological consideration is the stimulus duration used in clinical MP. The default 200-ms flash duration of the Nidek MP-1 is considerably longer than the brief flashes (typically 5–20 ms) or periodic flickering stimuli (5–30 Hz) commonly employed in conventional flash or steady-state VEP paradigms. As a result, classic time-locked VEP waveforms (e.g., P100) were not consistently observable in single trials. This necessitated the use of extended 600-ms EEG segments and a machine learning–based detection strategy rather than reliance on explicit waveform peak identification, as illustrated in the EEG preprocessing and segmentation framework (**Figure 4**). While this approach reduces interpretability—precluding attribution of responses to specific canonical VEP components—it enables stimulus detectability in conditions where traditional VEP analysis is not feasible.

Model performance revealed a clear trade-off between sensitivity and precision across trials (**Table 2**). When sensitivity was prioritized, precision decreased, resulting in increased false-positive detections, whereas higher precision was associated with missed stimuli. The overall performance pattern—characterized by high specificity and variable sensitivity—suggests a bias toward the majority class (non-stimulus segments), likely driven by class imbalance, low signal-to-noise ratio, and trial-specific noise characteristics. This variability is consistent with the small dataset and heterogeneous recording conditions. Future work incorporating cost-sensitive loss functions, adaptive decision thresholds, and class-balancing strategies may help mitigate this trade-off. Importantly, model training and testing were performed within individual trials, which may yield optimistic performance estimates due to shared noise properties. Cross-trial and cross-subject generalization was not evaluated and will be a critical focus of future validation studies. Accordingly, no claim is made that the present architecture is optimal; rather, it serves as a feasibility demonstration of stimulus detectability under constrained conditions.

Most existing approaches to objective microperimetry rely on spatially dense, cortically scaled stimuli and extensive averaging, which are not compatible with the non-standard, sparse, and long-duration stimuli used in clinical MP. The central question motivating this study was whether deep learning could detect subtle cortical responses to such constrained stimuli. By analyzing parieto-occipital EEG responses to isolated MP flashes without signal averaging, we demonstrate that detectable stimulus-related neural information persists even under substantial methodological limitations. Consistent with known visual cortex anatomy, bilateral occipital electrodes (O1/O2) provided the most robust performance across trials compared with parietal or combined montages (**Table 3**). We do not propose EEG-based detection of MP stimuli as a replacement for established mfVEP or mfPOP methodologies, which remain superior when optimized hardware, cortical scaling, and dense electrode arrays are available. Instead, our findings suggest that EEG may serve as a complementary modality capable of augmenting existing MP workflows.

A potential clinical implication of this approach is the automated registration of MP stimuli without reliance on patient button presses. By enabling more frequent presentation of catch or blind-spot stimuli without diverting patient attention, EEG-based stimulus detection could reduce fixation losses and improve test reliability, particularly in populations where subjective responses are unreliable, such as pediatric, elderly, or cognitively impaired patients. This conceptual workflow—from MP stimulus presentation to EEG-based detection and classification—is summarized in the experimental overview (**Figure 3**). These potential benefits, however, remain speculative and require rigorous validation.

Several limitations of this study warrant emphasis. First, the analysis relied on extended EEG segments and post hoc visualization of averaged signals to support hypothesis validation, rather than direct identification of canonical VEP components. As shown in the averaged waveform comparisons (**Figure 6**), stimulus and non-stimulus segments exhibit distinguishable low-frequency amplitude differences even in the absence of classic VEP morphology. Second, the within-trial training and testing strategy limits inference regarding generalizability to new trials or subjects. Third, the small sample size—two participants across 12 trials—precludes any claims regarding population-level performance. Finally, the absence of hardware-based synchronization introduced temporal uncertainty of up to 250 ms, substantially constraining detection fidelity.

**Figure 6:**
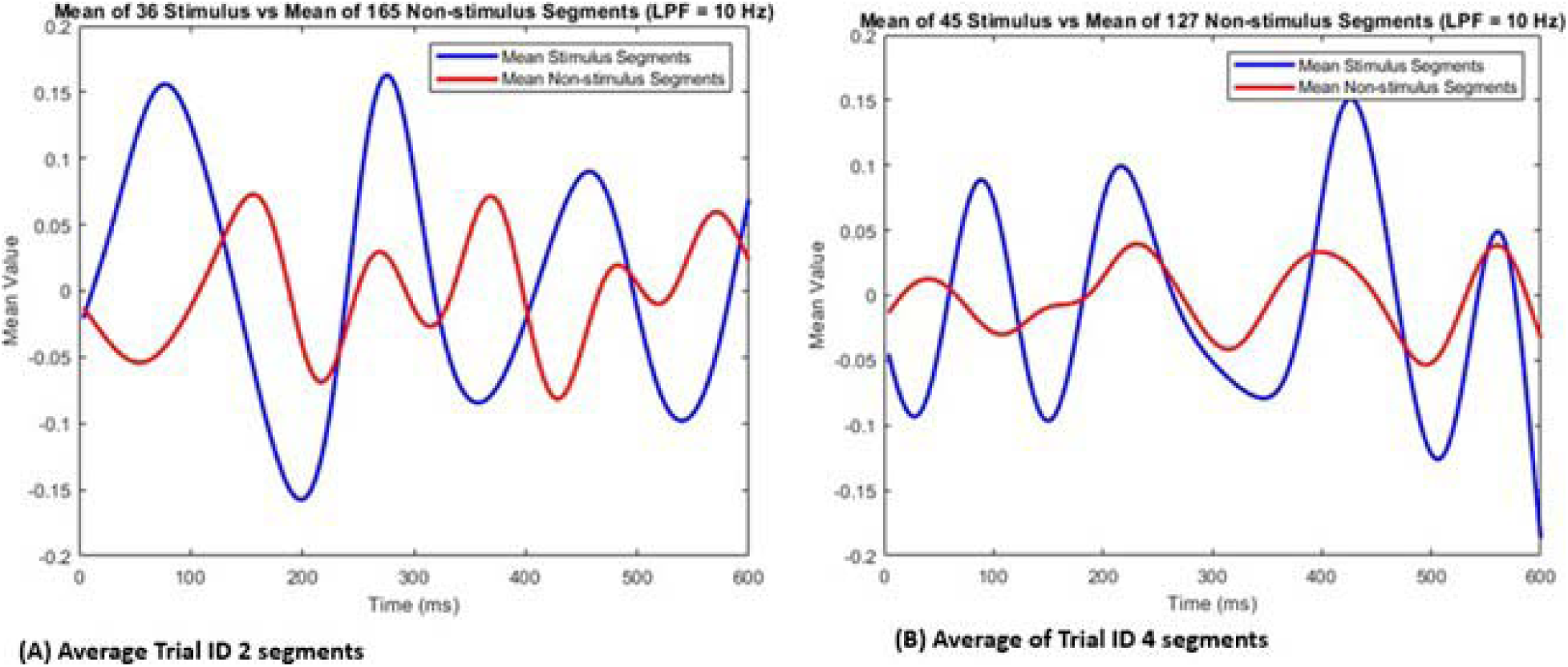
Averaging of all the segments to determine the existence of distinguishable VEP patterns from stimulus and non-stimulus segments (A) segments within trail ID 2 (B) segments within trail ID 4.

The observed performance improvements with higher-intensity stimuli and occipital electrode selection underscore the importance of precise synchronization and optimized signal acquisition. Future iterations of this work will incorporate hardware-level triggering to reduce timing uncertainty to the millisecond range and explore hybrid convolutional neural network–LSTM architectures to better capture spatiotemporal EEG features. With these refinements, real-time classification of stimulus-present and stimulus-absent EEG segments may become feasible, advancing the development of objective augmentation strategies for MP.

In summary, this study provides initial evidence that EEG signals can reflect single-flash stimuli in a microperimetry context, even under non-ideal acquisition conditions. While substantial technical and methodological advances are required before clinical translation, these findings support continued investigation into EEG-augmented microperimetry as a potential objective adjunct, bridging the gap between current subjective testing paradigms and fully objective functional assessment.

## Conclusion

This study provides initial evidence that EEG signals can reflect single-flash MP stimuli even in the absence of precise synchronization. While significant technical refinements are required before clinical translation, these findings support continued investigation into EEG-augmented MP as a potential objective adjunct, particularly for populations in whom subjective responses are unreliable.

## Data Availability

All data produced in the present study are available upon reasonable request to the authors.

## Ethical Approval

The Stanford institutional review board (IRB no. 68008) approved this study.

## Acknowledgments

None.

## Authors’ Contributions

YJS and MIA conceptualized the study. MASS, ASDN, KJ, TS and ZZF collected the data. MND analyzed the data. MND and ASDN wrote the first draft. All authors reviewed, edited, and approved the final draft.

## Abbreviations

ADAM: adaptive moment estimation
AMD: age-related macular degeneration
BiLSTM: bidirectional long short-term memory
BCVA: best-corrected visual acuity
CMR: common median reference
CNN: convolutional neural network
DA-VEP: dark-adapted visual evoked potentials
DME: diabetic macular edema
EEG: electroencephalography
FERG: focal electroretinography
GA: geographic atrophy
GUI: graphical user interface
HVF: Humphrey visual field
LSTM: long short-term memory
mfPOP: multifocal pupillographic objective perimetry
mfVEP: multifocal visual evoked potential
OCT: optical coherence tomography
PDF: portable document format
MP: microperimetry
SD: Stargardt disease
SD-OCT: spectral domain optical coherence tomography
SNR: signal-to-noise ratio
VEP: visual evoked potentials

## Appendix

